# Penalised neural network with weight correlation descent for predicting polygenic risk score

**DOI:** 10.1101/2023.05.23.23290438

**Authors:** Sun bin Kim, Joon Ho Kang, MyeongJae Cheon, Dong Jun Kim, Byung-Chul Lee

## Abstract

In this study, we developed a deep neural network (DNN) model with weight correlation descent (WCD) regularization to improve polygenic risk score predictions for complex diseases, specifically gender-specific cancers, using the UK Biobank dataset. Our DNN model with WCD outperformed both conventional PRS models and DNN models without WCD, demonstrating the importance of regularization techniques in enhancing model performance and capturing non-linear effects and interactions in genomic data. These findings contribute to a better understanding of genetic architecture, facilitating personalized interventions based on individual genetic profiles and ultimately benefiting patient care and health outcomes.

## Introduction

Genome-wide association studies (GWAS) have led to numerous scientific and biological discoveries by identifying a wide range of associations between common genetic variants and complex traits in populations^1^. Recent GWAS studies with increasingly larger sample sizes have uncovered more accurate and novel significant associations between various diseases or traits^1^. The probabilistic susceptibility of an individual to a disease, referred to as the polygenic risk score (PRS), can be estimated through GWAS results. PRS also allows us to understand genetic architecture and support clinical decisions, particularly in risk stratification, early disease detection, and prevention of common adult-onset conditions^2–5^.

In the early PRS development phase, unadjusted PRS were calculated using statistically significant variants from GWAS^6^. However, overfitting can still occur with small sample sizes or high number of variants^7–9^. Alternative approaches employ regularization models like BLUP^10^ and gBLUP^11^, but these can overestimate the effect size of correlated variants.

To address overestimation, linkage disequilibrium (LD) was applied. One method selects representative SNPs in LD blocks (P+T), while another uses LD panels, forming the basis of LDpred^12^, Lassosum^13^, and PRS-CS^14^. However, these methods require costly LD reference panels and may not accurately reflect real-world scenarios with non-linear effects and interactions.

Predicting polygenic risk using machine learning models can capture variant interactions and non-linear effects with fewer genetic architecture assumptions^15^. A breast cancer PRS model, which applied deep neural network (DNN)^15^, outperformed various machine learning models and conventional PRS models. Additionally, a study on Alzheimer’s disease PRS prediction employed DNN models^16^, which surpassed traditional PRS models and graph neural network (GNN) models. Despite DNN’s superior performance, overfitting can still occur, necessitating additional regularization in machine learning methods, including deep learning.

Regularization techniques are essential in deep learning to prevent overfitting and improve model generalization on unseen data. Common approaches include L1 and L2 regularization, dropout^17^, early stopping, weight decay, and batch normalization^18^. In addition, weight correlation descent (WCD)^19^ has been introduced as a regularization method for deep learning models. WCD further improves the model’s performance by preventing overfitting and guiding the model’s nodes to learn unique information from the input data. These methods, including WCD, help manage high-dimensional data and contribute to more accurate and generalizable models.

In this study, we exploited WCD to as a regularization method to a DNN model, testing it on real data from the UK Biobank^20,21^ to predict the genetic risk of gender-specific cancers, including breast cancer and prostate cancer. The DNN model outperformed conventional models, including P+T, PRSice, and PRS-CS in Nagelkerke R-square and the area under the receiver operating characteristic curve(ROC AUC). Importantly, the DNN model with WCD achieved significantly better performance than the plain DNN model.

The results demonstrate that the DNN model with WCD improves genetic risk predictions for complex diseases by accounting for non-linear effects, interactions, and preventing overfitting. The enhanced understanding of genetic architecture, facilitated by WCD, can inform personalized interventions based on individual genetic profiles.

## Results

### Overview of methods

We acknowledged that overfitting posed a significant challenge due to the presence of highly correlated genetic features within DNN models. To address this issue, we employed WCD as an appropriate regularization technique. WCD is designed to reduce the interdependence among nodes in a DNN model by guiding each node to train in a unique direction. This diversification of training directions allows the model to better capture the underlying patterns within the genetic data, ultimately enhancing its predictive capabilities.

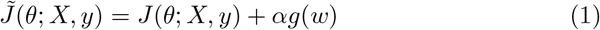

WCD accomplishes its objective by incorporating the cost of weight correlation into the model’s loss function, as illustrated in Equation eq. 1. In this equation, *J*(*θ*; *X, y*) represents the original loss function, while *g*(*w*) denotes the cost function associated with weight correlation. Furthermore, *α* serves as the coefficient determining the contribution of weight correlation during the training process. By integrating these components, WCD effectively reduces interdependence among nodes, enhancing the model’s stability and performance.

By doing so, we ensure that the weights of nodes in the l-th layer develop independently from each other, preventing any unnecessary influence among nodes. This approach not only mitigates the risk of overfitting but also contributes to improved stability and generalization in our DNN model.

### GWAS

In our study, we conducted a Genome-wide Association Study (GWAS) using the UK Biobank (UKBB) dataset to identify genetic associations with various traits. Two types of datasets were analyzed: the UKBB array data and an imputed UKBB dataset. Imputed datasets are generated by estimating genotypes at untyped loci based on known linkage disequilibrium (LD) information, which increases the resolution of the dataset. The number of samples used in GWAS and the number of SNPs are summarized in tbl. 1.

**Table 1:**
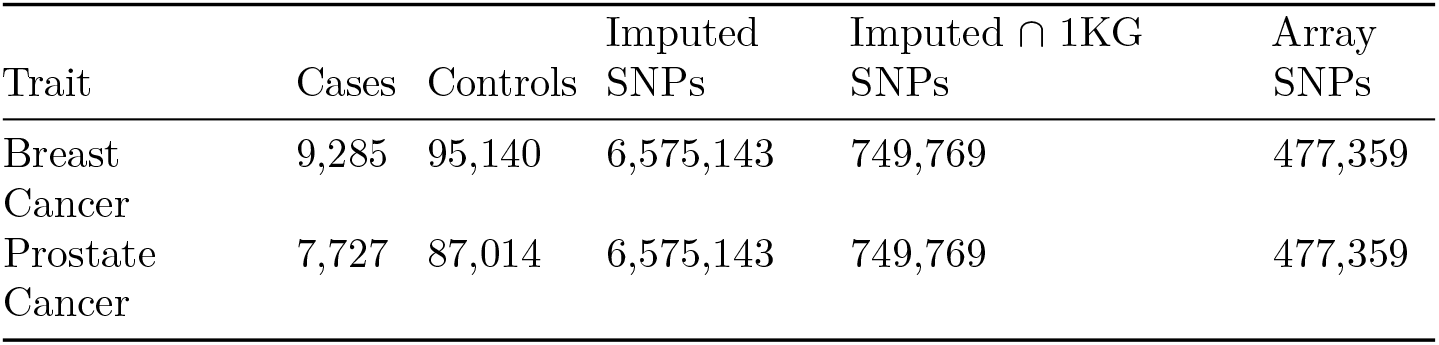
GWAS Information

To assess the quality and effectiveness of the GWAS, we relied on Manhattan plots and Q-Q plots. Manhattan plots are graphical representations of the statistical significance of genetic variants across the genome. These plots in fig. 1 revealed the presence of several significant genetic variants associated with the traits under investigation, confirming that the GWAS had successfully identified meaningful associations.

**Figure 1:**
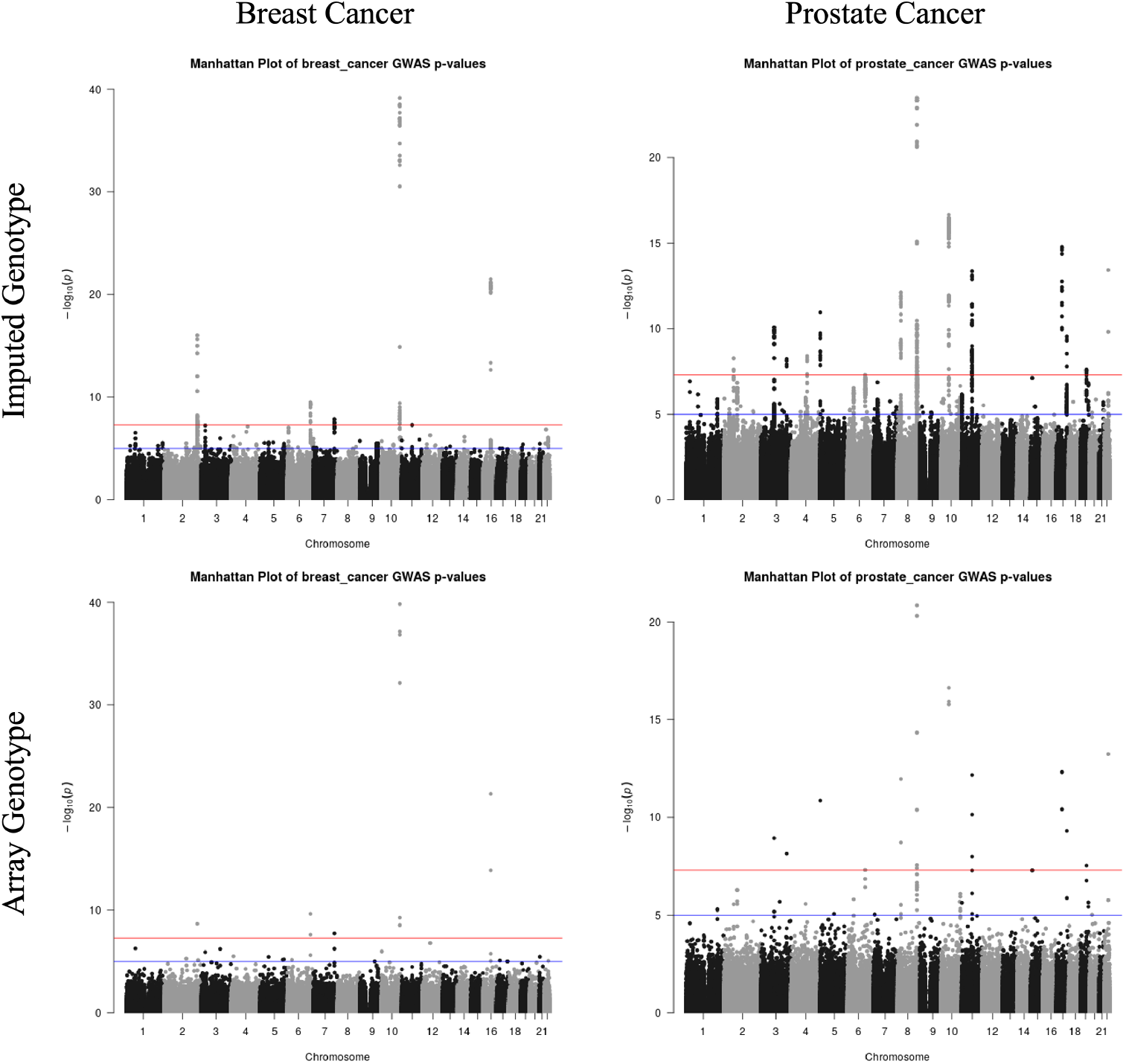
Manhattan Plot for the conduected GWAS

The imputed dataset, which incorporates LD information, demonstrated more significant variants in the same LD block. These additional variants were highly correlated with the significant variants found in the original array genotype data, suggesting that they are potentially capturing similar genetic effects.

Q-Q plots were used to compare the distribution of p-values obtained from the GWAS to the expected statistical distribution under the null hypothesis. The plots in fig. 2 showed a significant deviation of the observed p-values from the expected distribution, indicating that the GWAS had effectively identified true genetic associations with the traits.

**Figure 2:**
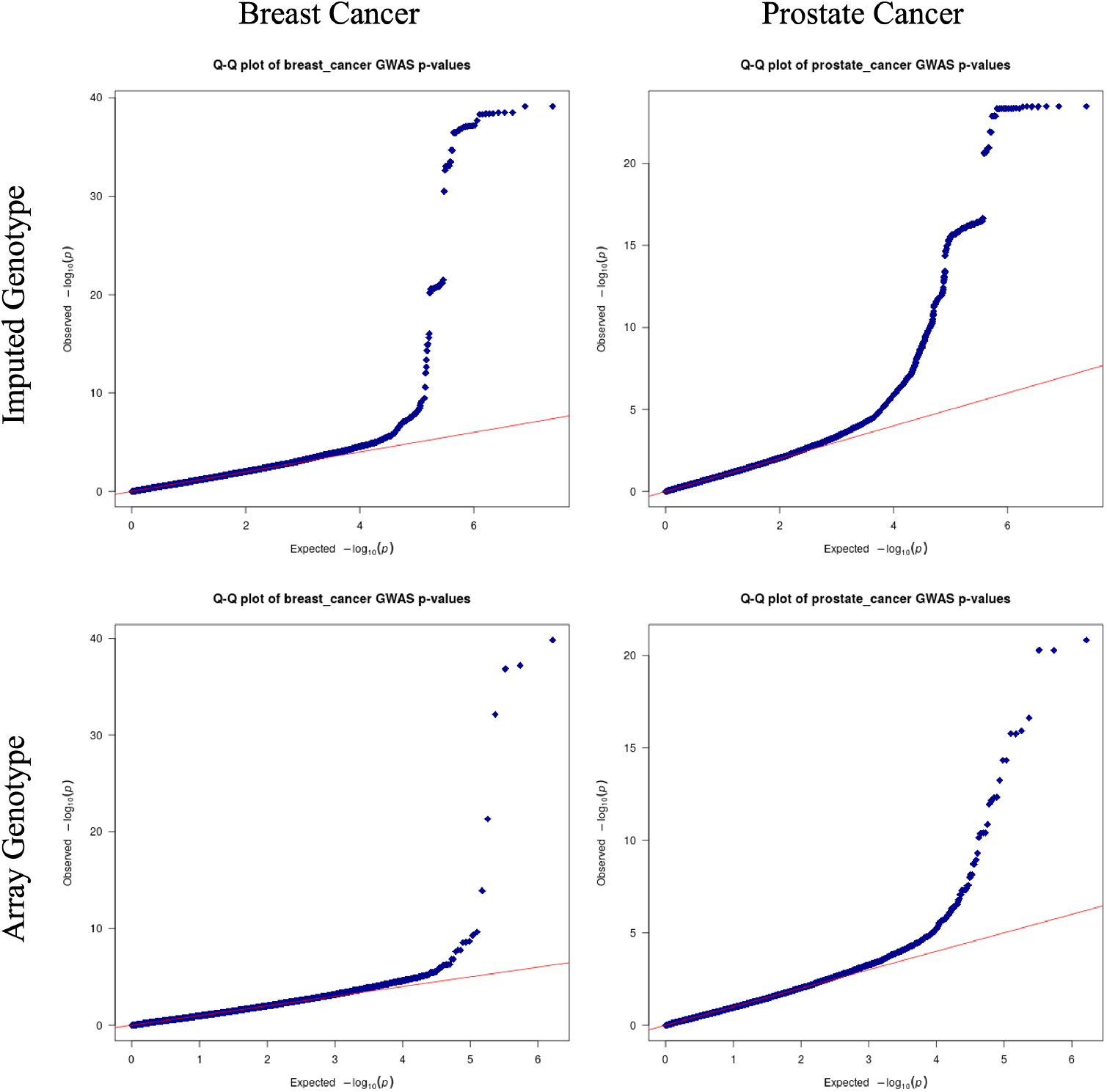
Q-Q plots for the conducted GWAS

It is worth noting that the figures for the imputed and array datasets were different. The imputed dataset’s Q-Q plot exhibited denser points, which can be attributed to the inclusion of additional genetic variants based on LD information. Overall, the results of our GWAS analysis using both the UKBB array data and imputed UKBB dataset demonstrate the successful identification of genetic associations with the traits of interest.

### Polygenic Risk Score

In this study, we aimed to explore the potential of penalised DNN models in predicting gender-specific cancers, including breast cancer and prostate cancer, by comparing their performance with conventional models. Our analysis included the baseline models P+T, PRSice, and PRS-CS, which used two LD panels (1KG and UKBB), as well as a DNN model and a DNN model with WCD as a regularization method. To evaluate the performance of these PRS models, we employed two widely accepted metrics: Nagelkerke R-square, ROC AUC and age-case incidence plots, which compare the top 5% of participants with the highest polygenic risk to the bottom 5% of the sample, yielded comparable results in terms of case incidence with respect to age. These findings are detailed in Supplementary Data 1.

For conventional models, we relied on imputed UKBB genotype data, as these models typically use summary statistics derived from imputed genotype data to build their predictive models. In contrast, for DNN models, we employed UKBB array genotype data during GWAS to reduce feature dimensionality and prevent the inclusion of features containing highly correlated variants with statistically significant associations to the trait of interest. We used GWAS results to select significant variants with a range of p-value thresholds for the DNN model.

Our results in fig. 3 demonstrated that the PRSice model had the lowest performance in both metrics for both breast and prostate cancer, with Nagelkerke R-square scores of 0.0079 and 0.01, respectively. The P+T model achieved slightly better Nagelkerke R-square and AUC scores than the PRSice model, with a 16% higher Nagelkerke R-square score for prostate cancer. The PRS-CS model outperformed both in terms of Nagelkerke R-square. Notably, PRS-CS with the 1KG LD panel had the highest score among the conventional models for breast cancer, with a 14% higher Nagelkerke R-square score compared to the P+T model. In contrast, PRS-CS with the UKBB panel achieved the highest Nagelkerke R-square score among linear models for prostate cancer, with a 40% higher score compared to the P+T model. The DNN models exhibited superior performance in predicting both breast and prostate cancer compared to all conventional models. The DNN model with WCD regularization achieved the highest scores among all models, including the plain DNN model, for both cancer types. Specifically, the DNN model with WCD achieved a 5.4% higher Nagelkerke R-square score for breast cancer and a 2.2% higher Nagelkerke R-square score for prostate cancer than the plain DNN model. In terms of AUC, the DNN model with WCD achieved slightly better performance than the plain DNN model for both cancer types.

**Figure 3:**
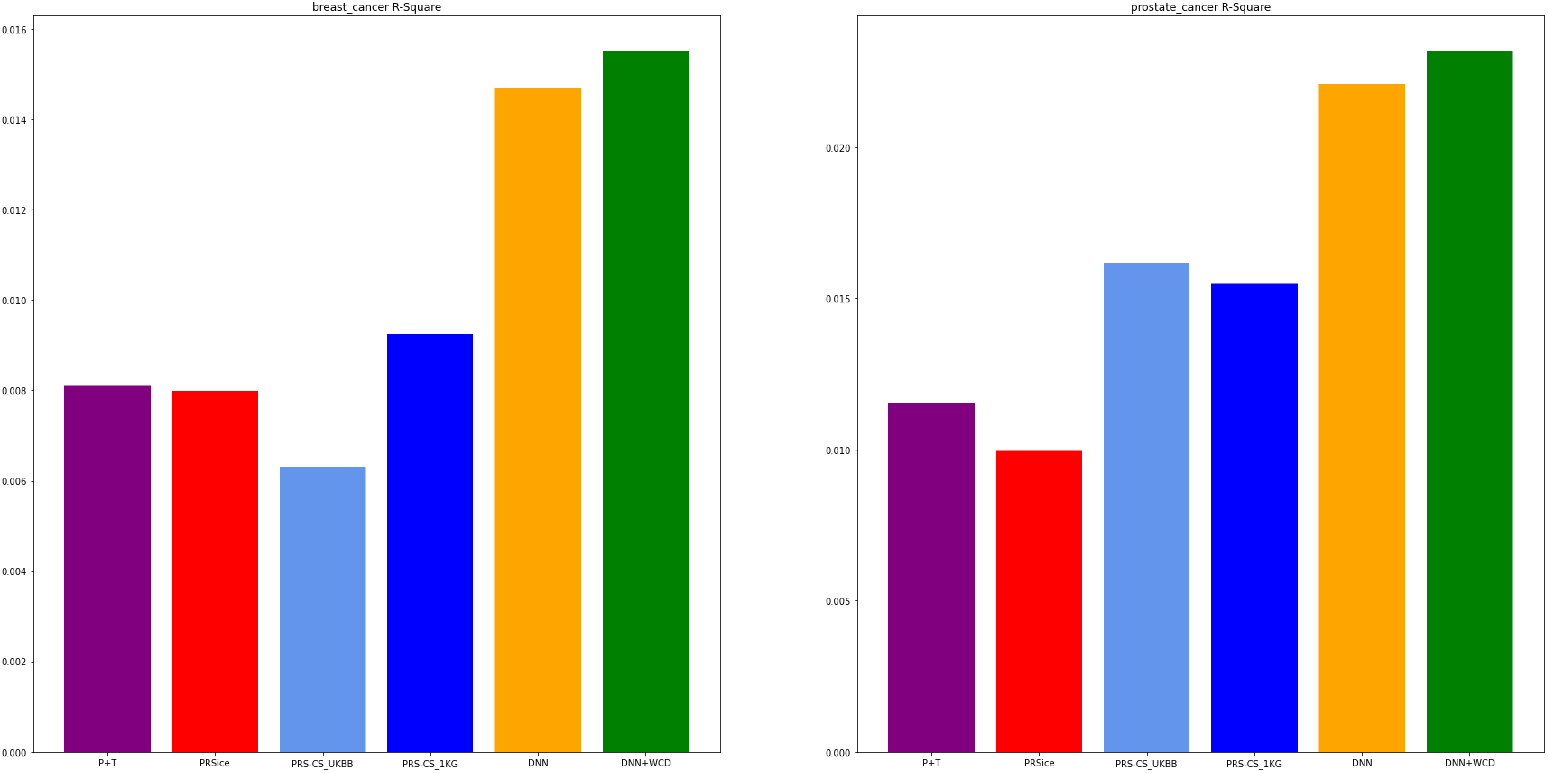
Nagelkerke R-Square Plot

In terms of AUC, a similar trend was observed for the AUC metric in fig. 4 as with Nagelkerke R-square, although the differences between the models were relatively smaller due to the different scales of scores. The PRSice model achieved the lowest AUC scores for both breast and prostate cancer. The P+T model followed the PRSice model with slightly higher AUC scores for both cancer types.For breast cancer, the PRS-CS model with the UKBB LD panel had the lowest AUC score, while the PRS-CS model with the 1KG LD panel achieved the highest AUC score among the conventional models. In the case of prostate cancer, the PRS-CS model with the UKBB LD panel achieved the highest AUC score, similar to the trend observed with Nagelkerke R-square. The PRS-CS model with the 1KG LD panel also performed well, but with a slightly lower AUC score. In predicting genetic risk for both breast and prostate cancer, the DNN model outperformed conventional models, including the PRS-CS model. Furthermore, the DNN model with WCD regularization demonstrated slightly better performance than the plain DNN model.

**Figure 4:**
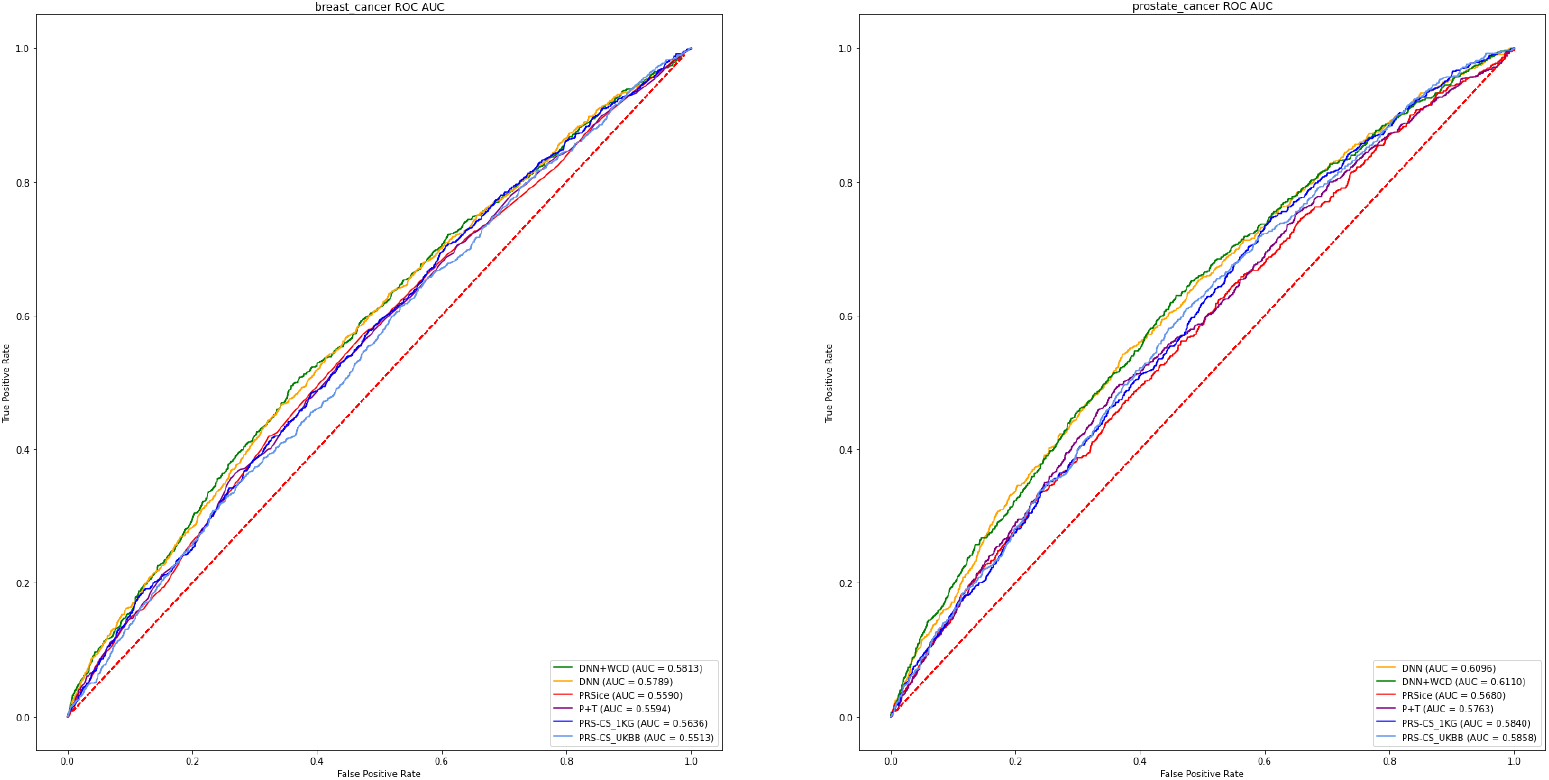
ROC AUC Plot

## Discussion

Polygenic prediction is a promising approach in clinical translation in genetics, as it uses genome-wide genetic markers to estimate an individual’s predisposition to complex human diseases or traits. This technique has the potential to improve clinical care by enabling targeted interventions based on a person’s genetic profile. The inherent complexity of genotype data, with its massive dimensions, poses a significant challenge when developing PRS models.

The aim of this study was to improve polygenic risk score predictions for complex diseases using a deep neural network (DNN) model with an appropriate regularization method, specifically weight correlation descent (WCD). We addressed the limitations of existing DNN models by constructing a DNN model capable of capturing non-linear effects and interactions in genomic data while avoiding distraction from noisy signals. Our key findings revealed that the DNN model with WCD outperformed existing PRS models in terms of Nagelkerke R-square and ROC AUC metrics, suggesting the presence of non-linear genetic architecture that cannot be well explained by linear models alone.

When comparing our results with previous studies^15,16^ that applied deep learning models to predict PRS, we found that while they focused on selecting significant SNPs using traditional methods, our research emphasized constructing DNN models with both explicit and implicit regularization techniques to enhance their performance. Our study highlighted the importance of regularization in PRS models and demonstrated the potential of DNN models with WCD for constructing PRS models using biobank-level databases, even when dealing with a relatively small number of cases.

Our study encountered several limitations that could impact the interpretation of the results. One such limitation was the reliance on GWAS-selected SNPs as input for the DNN model, which filters out SNPs lacking linear correlation to traits but may still have an effect. This constraint means our approach is unable to identify novel variants. Furthermore, GWAS might filter out certain combinations of SNPs that have a collective effect while not being significant individually. Selecting such combinations is computationally expensive, necessitating alternative solutions like machine learning or deep learning approaches. Additionally, the interpretation of DNN models is challenging, as popular XAI models like LIME^22^ and Grad-CAM^23^ only provide information about individual feature contributions, not the combination of features.

Another limitation is that DNN models require a large number of samples, making them unsuitable for diseases or traits with low prevalence or insufficient case samples. The high-dimensionality of genomic data and complex LD patterns hinder the model’s training, and learning LD information from samples for DNN models requires a relatively large number of samples. The dimensionality issue also leads to overfitting in PRS models using raw genotype data.

Future research should focus on improving the model’s ability to predict phenotypic probabilities with fewer samples through dimension reduction techniques or pre-training. For instance, if a pre-trained extractor can condense the input, predicting polygenic risk scores might require fewer samples compared to plain DNN models. As genetic variants have unique characteristics like smaller vocabularies, it is challenging to use pre-trained models from language or vision domains without modifications to address genetics problems. Developing model architectures and genome-specific training methods capable of understanding and learning the particular features of genomics data is essential to overcome these limitations and advance the field.

In conclusion, this study demonstrates the potential of a DNN model with WCD for improving polygenic risk score predictions for complex diseases by capturing non-linear effects and interactions in genomic data. By overcoming the limitations of existing approaches and focusing on regularization methods, this research contributes to the advancement of polygenic prediction and its clinical translation, ultimately benefiting patient care and health outcomes.

## Methods

### Participants

UK Biobank Axiom Array genotyped data^20^ was used to build PRS models for two cancers with high prevalence and heritability: breast cancer and prostate cancer. The UK Biobank gathered around 500,000 participants aged 37-73 years during recruitment from 2006 to 2010, conducting a genotyping and a surveying about baseline characteristics and so on. The participants were limited to white British individuals validated based on self-report and genetic data. The participants related with neither first-degree, second-degree, nor third-degree relatives were also restricted as well as whose genetic information was not available or with presence of aneuploidy.

UK Biobank was given written informed consent by all participants to access their data and samples for research purposes. Ethical approval to collect and use participants’ data of UK Biobank has been acquired by the North West Multicentre Research Ethics Committee, the National Information Governance Board for Health & Social Care, and the Community Health Index Advisory Group. All relevant guidelines and regulation were followed while conducting this study.

### Ascertainment of Cancer Incidence

Disease outcomes were identified using hospital episode statistics. To classify cases and controls of cancers, we extracted International Classification of Diseases (ICD) codes from hospital admissions (versions ninth and tenth) and self-report disease data, following guidance from previous literature^21^. Cases and controls for diseases were identified according to criteria referenced from existing studies^24,25^.

Breast cancer cases were defined as women with a malignant neoplasm of the breast identified by ICD9, ICD10, or self-report data. Similarly, men with a malignant neoplasm of the prostate, as determined by either ICD9, ICD10, or self-report data, were classified as prostate cancer cases. Controls were defined as individuals without any cancer-related signals, with females considered as controls for breast cancer and males for prostate cancer, respectively. In this study, the total number of breast cancer incidences used was 104,425, consisting of 9,286 cases and 95,139 controls. Additionally, the prostate cancer dataset comprised 7,487 cases and 87,254 controls, totaling 94,741 individuals.

### Quality Control

Before calculating the PRS, genotyped data must undergo quality control to reduce genotyping and imputation errors. The quality control procedure for both genotyped and imputed genotype data includes the following steps: removal of genetic variants (1) with missing call rates exceeding 0.05, (2) with allele frequency below 0.01, or (3) with Hardy-Weinberg equilibrium exact test p-values below 1e-10. We also removed ambiguous SNPs and deduplicated variants based on their position, retaining only SNPs present in the genotype data of the UK Biobank and used summary statistics. Additionally, for imputed genotype data, variants with INFO scores below 0.7 were excluded.

### GWAS

To select statistically significant SNPs from a genome-wide range of variants and estimate their effect sizes, we used summary statistics of GWAS obtained from partial samples of the UK Biobank. These GWAS and the number of SNPs are summarized in tbl. 1. To validate and test the PRS models, the dataset was split into three subsets: train, validation, and test. First, we stratified the dataset based on case-control information. Second, we divided it into a training set and a test set in a 9:1 ratio. Lastly, the divided training set was further split in an 8:2 ratio and used as a training set and a validation set. The training set was used to conduct GWAS, preventing data leakage.

### Baseline Models

We considered three of the most popular PRS approaches: P+T, PRSice^26^, and PRS-CS^14^ as baseline models. These conventional PRS models were constructed using imputed genotype data from the UK Biobank Axiom Array. PRS was calculated as a linear combination of each variant’s beta coefficient (effect size) and the number of alleles present at that position under an additive genetic architecture model. Specifically, PRS for the i-th individual is calculated as eq. 2, where M is the number of genetic markers used for PRS, *X*_*ij*_ is the number of alleles for the j-th SNP of the i-th individual, and 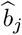 is the projected effect size of the j-th SNP.

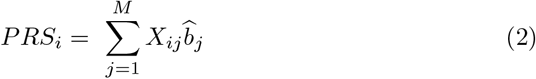

The P+T method aims to find valid causal variants associated with traits or diseases by pruning SNPs correlated with LD and applying a p-value cutoff to remove variants without sufficient statistical significance. In this study, the P+T model was implemented with PLINK^27^, pruning with *r*^2^ ≥0.1, which is the correlation between SNPs having a physical distance (called window size) smaller than 250 kb, and then thresholding with the p-value of 5e-8, according to the standard^28^. The PRS of the P+T model was calculated as eq. 2, using the selected SNP set and the beta coefficient of GWAS summary statistics without any modification.

As mentioned above, since static p-value and pruning parameters might not be optimal, the PRSice model attempts to find optimal parameters used in the P+T procedure by validating them with metrics in an iterative way. We reproduced the PRSice model using a software package published on GitHub by Choi, S.W. & O’Reilly, P.F^26^, with default parameter settings. Specifically, during the pruning step, we fixed the window size at 250 kb and *R*^2^ criteria at 0.1. In the thresholding stage, less significant SNPs were removed using p-values as listed below. We considered threshold *P*_*T*_ values from 5e-8 (the standard) to 1 (which uses all SNPs, called the full model) with an interval of 5e-5 in this paper. The polygenic score is then calculated as the sum of the remaining, largely independent SNPs with GWAS association p-values below a threshold *P*_*T*_, weighted by their marginal effect size estimates. Ultimately, the *P*_*T*_ value with the highest prediction accuracy in a validation dataset is applied, and the model’s performance is evaluated using an independent testing set.

The PRS-CS model estimates PRS by not only using the beta coefficients of GWAS but also modifying the beta coefficients through a method called continuous shrinkage. This method transforms the distribution of the beta coefficients using a predefined prior distribution and the LD reference. The PRS-CS model considers the following phenotype model:

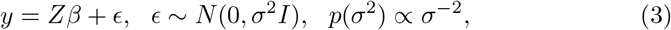

The prior distribution is based on global-local scale mixtures of normals as shown in eq. 4, where *σ*^2^ is the variance of *β*_*j*_, N is the number of samples, *ϕ* is a global scaling parameter that affects the effect size of all variants, *ψ*_*j*_ is a local and marker-specific parameter, and g is an absolutely continuous mixing density function.

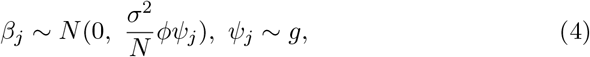

The posterior mean of *β* is

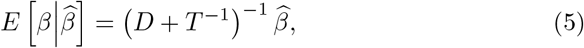

where *T* = *diag* {*ϕψ*_1_, *ϕψ*_2_, …, *ϕψ*_*M*_} is a diagonal matrix and *D* = *Z*^*T*^ *Z/N* is the LD matrix, with Z being an *N*× *M* standardized genotypes matrix. The local shrinkage parameter *ψ*_*j*_ was assigned an independent gamma-gamma prior as

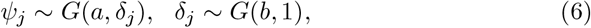

The PRS-CS was implemented using the software package published on GitHub by Ge, T. et al.^14^.

### Deep Neural Network

In this study, we propose a deep neural network (DNN) model to predict genetic risk using summary statistics for selecting a significant SNP set. A variety of DNN architectures were trained and validated using grid search, which identifies optimal hyperparameters and an architecture for the DNN model with the highest validation score. The Leaky Rectified Linear Unit activation function^29^ (Leaky ReLU) was used for every hidden layer neuron as follows:

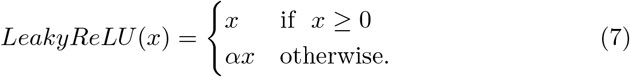

On the output layer, the logistic function was applied as:

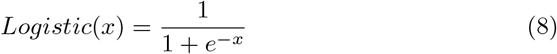

The loss function was computed using the binary cross-entropy function as:

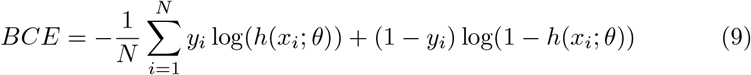

Here, *y* ∈{0, 1} is the prediction target, with 1 for cases and 0 for controls. *h*(*x*_*i*_; *θ*) ∈ [0, 1] is the predictive probability from the model for the target, given the input *x*_*i*_ and its parameters *θ*. The estimated probability is considered as the PRS projected by the DNN. The adaptive learning rate optimization algorithm, Adam optimizer^30^, was employed as the DNN model optimizer. The initial learning rate was set to 1e-3. Dropout^17^ was applied to reduce the probability of overfitting with a rate of 0.5. Additionally, Batch Normalization (BN)^18^ was used to enhance the training process of DNN models by reducing internal covariate shift. When used, Dropout and BN were applied to all hidden layers of the DNN models.

The grid search algorithm experimented with a list of DNN models with varying depth and width of hidden layers. The depth ranged from 1 to 4, and the width was determined by dividing the number of outputs from the previous layer by an integer from 1 to 4, which may differ from the number of selected SNPs for input. The hyperparameter combinations tested in grid search experiments also included models with and without regularization: BN and Dropout. Early stopping is employed to expedite the grid search process by halting training when performance improvement plateaus, thus preventing excessive computation time.

The SNP set, which served as the input for the DNN model, was selected based on p-values within a specific range. Since the scale of the p-value for summary statistics depends on the dataset used in GWAS, a list of p-values was handcrafted and chosen for each study and disease.

The DNN models were trained and validated similarly to conventional models. The performance of the DNN models was validated using the average ROC AUC of test sets. The best model for each p-value threshold was used as the base model for the stacking method, which will be discussed in the following section. Unlike the PRSice model, the DNN model does not use its beta coefficient, so internal GWAS summary statistics can be used to select the SNP set without the risk of overfitting. The grid search and DNN models were implemented using Python3 and PyTorch.

### Weight Correlation Descent (WCD)

The correlation between features can likely cause overfitting, especially in nonlinear models such as DNNs. To reduce the risk of overfitting caused by linkage disequilibrium (LD), weight correlation descent (WCD)^19^ can be helpful. WCD is a method to regularize the learning process by adding an additional term to the loss function for the model, as shown in eq. 10. Here, 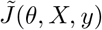 represents the regularized loss function, *J*(*θ*; *X, y*) is the loss function before applying WCD. The term *g*(*w*) represents the measure of correlation between the weights of each node in the DNN. *α* is the coefficient to adjust the effect size of WCD.

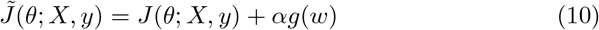

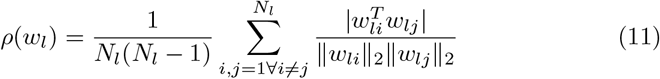

The weight correlation for each layer of the DNN is calculated using eq. 11, and *ρ*(*w*_*l*_) is used to obtain *g*(*w*_*l*_), which is the weight correlation loss for each layer, as shown in eq. 12. Then, *g*(*w*) can be achieved by simply summing the weight correlation loss for each layer: *g*(*w*) = Σ_*l*_ *g*(*w*_*l*_).

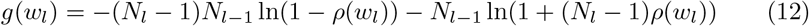

## Data Availability

This study was conducted using the resources of UK Biobank: www.ukbiobank.ac.uk (application ID: 72128). The PRS-CS algorithm was used as baseline model, implemented in python3, which is available at https://github.com/getian107/PRScs. Likewise, the PRSice algorithm was used as baseline model, contructed with R, which is available at https://github.com/choishingwan/PRSice. Most of genotype data preprocessing was done with PLINK2, which is available at https://www.cog-genomics.org/plink/2.0/.

https://github.com/getian107/PRScs

https://github.com/choishingwan/PRSice

https://www.cog-genomics.org/plink/2.0/

