## Supplementary Data 1 for "Penalised neural network with weight correlation descent for predicting polygenic risk score"

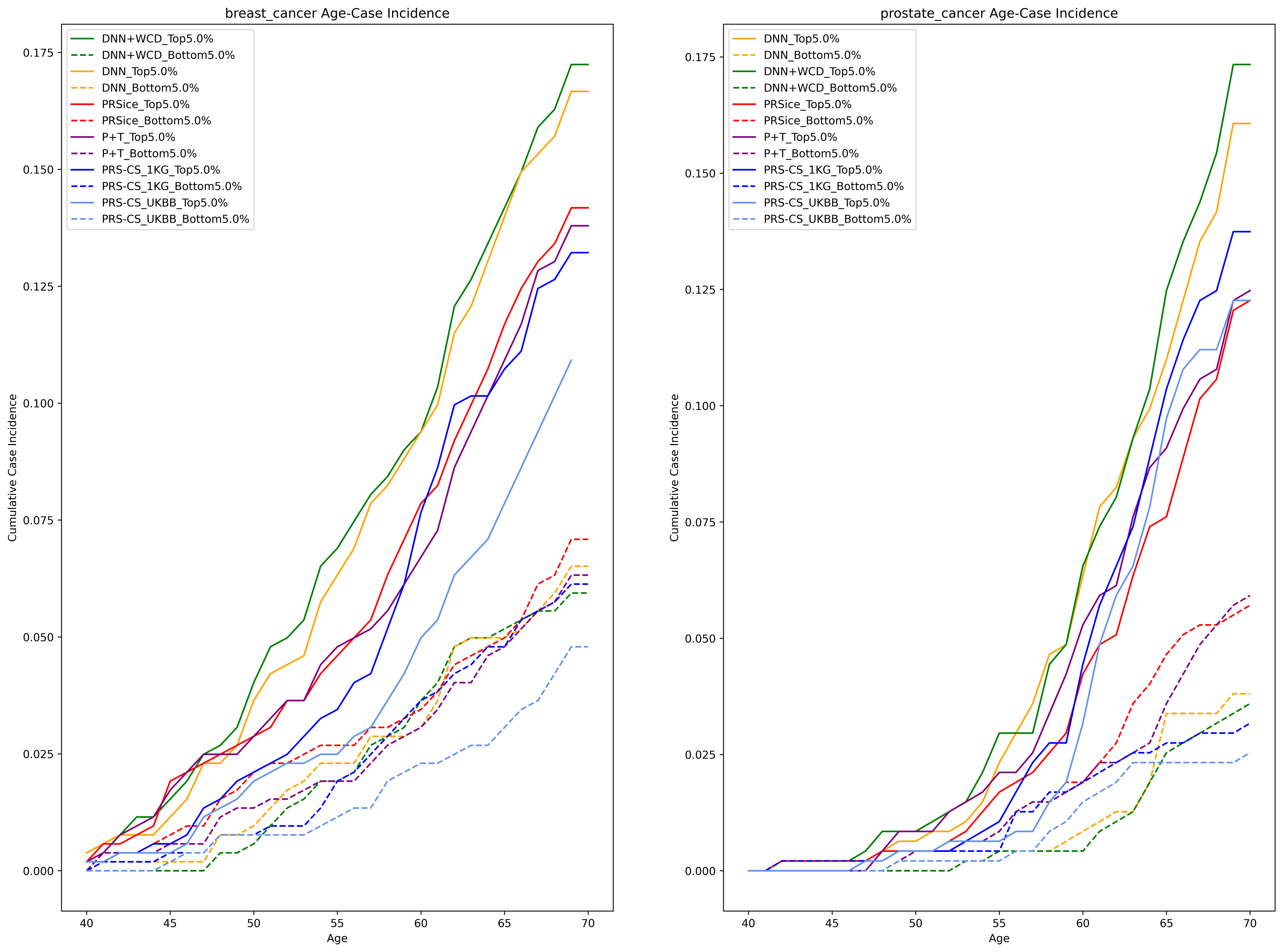

Figure S1: Age-case incidence Plot

Fig S1. displays a graph illustrating the case incidence per age from 40 to approximately 70, which represents the age range of case and control samples in the UK Biobank database, predicting whether they are cases or controls for two gender-specific cancer: breast cancer and prostate cancer. For breast cancer, we can see DNN models are relatively good at stratifying the cases, which is about 17.5% of cases existed in people with top 5% PRS. Moreover, DNN models stratified the control samples with the lower PRS, which is bottom 5%, including slightly more than 5% of cases. This is the lowest control ratio except for the PRS-CS with UKBB LD panel, which does not stratify well. For prostate cancer, similar trend was seen as the breast cancer, while DNN with WCD stratify the best for cases, PRS-CS model is the best model stratifying the control.
